# Feasibility of multicomponent exercise training with beat-accentuated music among community-dwelling older adults with mild-to-moderate cognitive decline

**DOI:** 10.1101/2023.04.30.23289323

**Authors:** Kyoung Shin Park, Lake Buseth, Jiyeong Hong, Jennifer L. Etnier

**Author notes:** Corresponding Author at Coleman 239, 1408 Walker Ave, Greensboro, NC 27412, USA. Disclosure statement: The authors report there are no competing interests to declare. Trial registration: ClinicalTrials.gov ID NCT05462977. Registered July 18, 2022.

## Abstract

**Objectives:** This study explored the feasibility and preliminary efficacy of a music-based, multicomponent exercise intervention among community-dwelling older adults with mild-to-moderate cognitive impairment.

**Methods:** 13 older adults aged 85±9 years with mild-to-moderate cognitive impairment completed multicomponent exercise training for 20 weeks at an independent living facility. Participants received aerobic, resistance, and balance training paired with beat-accentuated music stimulation (BMS). Participants’ adherence to the training was tracked down and their cognitive and physical functioning and quality of life were assessed at pre- and post-test.

**Results:** 13 participants attended an average of 4.6 days/week over 20 weeks and reported high satisfaction with the intervention (90.6%). Participants showed significant improvement in global cognition, cognitive processing speed, and walking endurance/aerobic fitness at post-test.

**Conclusions:** These findings support the feasibility of music-based, multicomponent exercise training for older adults in an independent living facility and set the stage for future studies to test the efficacy of music on physical activity and ensuing health outcomes.

**Clinical Implications:** Music-based, multicomponent exercise training can be beneficial for community-dwelling older adults with mild-to-moderate cognitive decline. BMS can be combined with exercise training to manipulate exercise tempo and may provide a source of motivation to help older adults adhere to exercise.

## Introduction

It is estimated that 13.8 million US adults will be living with Alzheimer’s disease (AD) by 2050, which will result in $1.1 trillion in health care costs (Alzheimer’s Association, 2020). With no curative treatment currently available, lifestyle interventions reducing risk factors are imperative to prevent or delay the onset of AD (Livingston et al., 2017). This is an important direction because delaying the onset of AD by 5 years could lower the prevalence of the disease by 42% and reduce the health care costs by $367 billion (Alzheimer’s Association, 2020). Notably, about one-third of AD cases worldwide are related to modifiable risk factors; the largest proportion of cases in the US is attributable to the lack of physical activity (PA) (Norton et al., 2014). Hence, it is important to promote PA in the aging population to protect against the progression of cognitive decline and dementia.

The global and national PA guidelines (PAG) prescribe older adults to regularly engage in moderate-intensity aerobic exercise for 150-300 min/week as well as resistance and balance training for 2-3 times/week (Bull et al., 2020; Piercy et al., 2018). However, current population data show that only 9-18% of US older adults adhere to the PAG (Bennie et al., 2019; Clarke et al., May 2017). This trend is untoward especially given the meta-analytic evidence that PA interventions with both aerobic and resistance training have shown greater benefits for older adults’ cognitive health compared with aerobic training only (Colcombe & Kramer, 2003). Despite the cumulative evidence supporting the benefits of exercise training for cognitive and brain health, the rate of citizens maintaining regular exercise is lower than desired (Kramer & Colcombe, 2018). Thus, a new approach is needed to promote adherence to the PAG in the growing number of older adults.

Our lab conceptualized a theory that music could be a motivational stimulant to promote regular PA (Park et al., 2023). Consistent with this perspective, a meta-analysis revealed that listening to music prior to or during acute exercise bouts increases positive affective valence (feeling good versus bad; *g* = 0.48, CI [0.39, 0.56]), reduces ratings of perceived exertion (RPE; *g* = 0.22, CI [0.14, 0.30]), enhances physical performance (*g* = 0.31, CI [0.25, 0.36]), and improves oxygen utilization efficiency (VO2max; *g* = 0.15, CI [0.02, 0.27]) among healthy adults (Terry et al., 2020). These findings support the notion that music helps exercise bouts to be perceived as more joyous, less arduous, and more energizing. Given that a negative affective response to exercise is a key barrier to PA whereas enjoyment is an important motivator of PA among older adults (Buman et al., 2010; Gray et al., 2016) and that humans are predisposed to take pleasure in moving in sync with music (Zentner & Eerola, 2010), exercising with music may help older adults engage in regular PA. However, it is important to note that not all music is effective for promoting regular PA.

Our lab conceptualized a theory that music could be a motivational stimulant to acute and long-term PA (Park et al., 2023). We further demonstrated increased positive affect and reduced RPE by exercising with Beat-accentuated Music Stimulation (BMS) in an acute phase compared with exercising without music among community-dwelling older adults (Park et al., 2022). As a form of rhythmic auditory stimulation, BMS refers to pulsed, tempo-synchronous music stimuli embedded with sonically enhanced beats to facilitate rhythmic body movement (Thaut et al., 2016). BMS has been employed in cardiac rehabilitation (Alter et al., 2015) and gait rehabilitation in people with Parkinson’s disease (Benoit et al., 2014; McIntosh et al., 1997; Thaut et al., 1996) or stroke (Prassas et al., 1997; Thaut et al., 1997). Alter et al. (2015) conducted a randomized controlled trial (RCT) and found that self-directed walking-for-exercise in sync with BMS led to nearly twofold increases in accelerometer-measured weekly volume of PA at all intensities and in caloric expenditure over 3 months among midlife-to-older adults in a home-based cardiac rehab program relative to the same exercise program with beat-unaccented, tempo-synchronous music or without music. These findings suggest that the distinctive combination of music with accented beats can dramatically increase PA above and beyond beat-unaccented music. However, to our knowledge, no study to date has applied BMS to multicomponent exercise training among older adults with cognitive decline.

We conducted a single-arm intervention trial to test the feasibility, acceptability, and preliminary efficacy of a 20-week BMS-based multicomponent exercise training program for community-dwelling older adults with mild-to-moderate cognitive impairment. Outcomes of interest were adherence to and overall satisfaction with the intervention as well as changes in cognitive and physical functioning and health-related quality of life (QoL). Our approach to prevent or delay the onset of AD or other dementias through multicomponent exercise training could be particularly urgent among older adults in early stages of cognitive decline. Given the low rate of PA among older adults in the US and the strong association between low PA and the prevalence of AD, developing and implementing a novel PA intervention for older adults will have implications for dementia prevention. This preliminary study will set the stage for an RCT to fully test the efficacy of BMS-based exercise training in the growing aging population at risk of dementia.

## Methods

### Participants

16 community-dwelling older adults (9 females) who were 86.2 ± 8.6 years old (*M* ± *SD*), previously low-active (< 60 min/week of exercise, determined by self-report and confirmed by facility staff), and with mild-to-moderate cognitive impairment (determined by Montreal Cognitive Assessment [MoCA]) were recruited from an independent senior living facility. Eligibility screening was conducted in person by appointment in a designated room at the senior living facility. Individuals were ineligible if they were incapable of walking, unable to hear verbal instructions and music, physically active (> 90 min/week of exercise), had severe cognitive impairment (determined by MoCA total score < 11; Nasreddine et al., 2005), or had anxiety or depression (determined by a single-item on the EuroQol Health Questionnaire [EQ-5D-5L] ≥ 4) (Herdman et al., 2011). Participants who could walk with or without an assistive device (walker, cane, etc.) or who could hear using hearing aids were eligible for the study. Participants also completed the Physical Activity Readiness Questionnaire for Everyone (Warburton et al., 2011) to determine if they had an ongoing medical condition that might put them at risk by engaging in moderate intensity exercise. Through this screening procedure, we excluded 1 individual with advanced Parkinson’s disease, 1 individual with post-stroke hemiparesis, and 2 individuals with MoCA scores < 11.

We were able to include participants with mild Parkinson’s disease, arthritis, or osteoporosis, taking medications to manage heart conditions and/or blood pressure, or using a walker for ambulation. These individuals were instructed to exercise at a light intensity and/or in a seated position within the training protocol. Eligible participants were provided with the study procedure and completed a Brief Informed Consent Test (Buckles et al., 2003) through which we confirmed their ability to understand the study information. All participants provided a written informed consent on their own. Among 16 enrolees, 3 withdrew participation due to unexpected lower body injuries or visual impairment (unrelated to the intervention) and thus 13 participants (7 females) with MoCA score ranging from 16 to 25 (*M* = 20.38, *SD* = 2.98) completed the intervention and were included in data analysis.

### Music-based multicomponent exercise intervention

All participants were provided with music-based multicomponent group exercise training at 9:30 AM for 30-35 min/day, 6 days/week over 20 weeks in a designated room at the senior living facility. The exercise goal for all participants was to attend the group exercise training > 3 days per week. The exercise program was open to all members in the residential facility and thus a few additional individuals often attended without participating in the study, but adherence was only tracked for study participants. To be consistent with the PAG, the group exercise program consisted of a dynamic warm-up (5 min), aerobic training (15 min), resistance and balance training (10 min), and cool-down stretches (5 min). See Table 2 for an overview of the exercise program. Most exercises were chair-assisted and thus adaptable across fitness levels and were safely implemented for participants with fall risks. Participants were instructed to exercise in a standing position with at least one hand holding a chair or in a seated position. The exercise program was developed and consistently delivered by two researchers along with three staff members in the facility, who were all CPR-certified and experienced exercise instructors. Participants were instructed to exercise at light-to-moderate intensity based on the Borg RPE Category-Ratio scale (Borg, 1982) which was posted in the exercise room.

Participants were trained to exercise in sync with the tempo of a BMS playlist that was made of 35 music excerpts, which were played in a randomized order. Exercise pace and the music tempo were incrementally increased by 5 beats per min (BPM) every 5 weeks during the intervention, from 85 BPM to 100 BPM. Participants were not asked to use BMS for their self-directed walking outdoors for safety reasons.

### Beat-accentuated Music Stimuli

All music excerpts in the BMS playlist were slow-to-medium tempo country and pop songs that were rigorously chosen by the author (JH), a trained musician and certified music production specialist, in consideration of participants’ music preference initially surveyed as well as their unchanging tempos and a clearly discernible rhythm in 2/4 or 4/4 meter. We then sonically enhanced each quarter note beat by adding lower- and/or higher-frequency drum sounds (kick-drum, snares, hihats, and rides) to correspond with one paced step or muscle contraction when exercising. The beats were added as a secondary track and recorded concurrently with the original music, using musical instrument digital interface (MIDI) keyboard-drum instruments (Pro Tools 2021, Avid Technology Inc., Burlington, MA), in a similar manner with Alter et al. (2015). Our intention was to implement beat accentuation at frequencies and volumes just beyond minimal detection levels without detracting from the authenticity of the original music. The tempo of music excerpts was adjusted without damaging the harmony or pitch via open-source sound-editing software (Audacity 3.0.4, The Audacity Team, available at audacityteam.org).

### Procedure

Individuals who expressed interest were scheduled for the screening by appointment in a designated area at the senior living facility. After screening, eligible participants completed pre-test to assess cognitive and physical functioning and health-related QoL. The total testing procedure was completed within an hour. Testing was conducted by research staff with the aid of facility staff. After a month of pre-testing all participants, the 5-month exercise intervention started and their adherence to the intervention was tracked down. After the intervention, participants were scheduled for post-test to assess the same outcome measures with the pre-test. At post-test, participants’ satisfaction with the intervention was also assessed.

### Outcome measures

Adherence to the intervention was tracked during the intervention through a sign-up sheet that was self-reported by participants and confirmed by the exercise instructor after every session. Participants’ satisfaction with the intervention was assessed using the Client Satisfaction Questionnaire (CSQ-8), an 8-item 4-point Likert scale with the total possible score ranging from 4 to 32 and a higher score indicating greater satisfaction (Larsen et al., 1979).

General cognitive functioning was assessed using the MoCA, a widely used test to access memory, executive function, and other symptoms of cognitive decline (Nasreddine et al., 2005). Version 8.1 and 8.2 of the MoCA was used at pre- and post-test, respectively. Test scores were calculated based on pre-established algorithms to obtain the MoCA total score and memory index score (MoCA-MIS) (Julayanont et al., 2014).

Inhibitory control and cognitive processing speed was assessed using the Flanker Inhibitory Control and Attention Test and the Pattern Comparison Processing Speed Test in the NIH Toolbox Cognition Battery (2022 Toolbox Assessments, Inc., available at nihtoolbox.org), The Flanker Test requires participants to indicate the left– right orientation of an arrow stimulus presented centrally while inhibiting attention to the potentially incongruent arrow stimuli surrounding the central stimulus. Accuracy and reaction time on the incongruent versus congruent items serve as measures of inhibitory control (Zelazo et al., 2013). The Pattern Comparison Processing Speed Test asks participants quickly identify whether two images are the same or not and the number of correct items completed in 90 sec is counted (Carlozzi et al., 2013). Both NIH Toolbox tests were validated in young-to-older adults aged 18–65 years (Weintraub et al., 2014) and oldest older adults aged 85–99 years (Nolin et al., 2022).

Health-related QoL was self-reported through the 5-item EQ-5D-5L (Herdman et al., 2011). Validated in older adults with multimorbidity (Bhadhuri et al., 2020), the EQ-5D-5L is based on descriptions of self-perceived health based on 5 dimensions: mobility, self-care, usual activities, pain/discomfort, and anxiety/depression. Each dimension has 5 response options corresponding to no problems, slight problems, moderate problems, severe problems, and incapability. Participants also reported their overall health on the day on a hash-marked, vertical visual analogue scale (EQ-VAS) marked from 0 (the worst health you can imagine) to 100 (the best health you can imagine).

Physical functioning was assessed through the Timed Up and Go (TUG), 4-Stage Balance Test (4SBT), 6-Minute Walk Test (6MWT), and 30-Second Chair Stand Test (30SCST) performed in that order. The TUG, 4SBT, and 30SCST are part of the CDC’s STEADI toolkit for the assessment of fall risks in older adults (Nithman & Vincenzo, 2019). The 6MWT is found to be a reliable and valid measure of physical endurance and aerobic fitness in older adults (Mänttäri et al., 2018; Rikli & Jones, 1998). We measured the duration to complete a TUG trial (after a practice trial), the sum of durations to maintain the posture required in the first, second, and third stage of the 4SBT, the total distance walked for 6 min (6MWT), and the reps of sit-to-stand maneuvers completed in 30 sec (30SCST). Some participants completed the tests using a walker as needed, consistently across the pre- and post-test.

### Data analysis

All statistical analyses were conducted with R 4.2.2 (R Core Team, 2022). Normality of the data was first checked with descriptive statistics and the Shapiro-Wilk test. For the outcome variables showing normal distribution, we conducted student’s paired-sample t-tests to examine the pre- and post-test differences with alpha (α) at < .05 for two-sided tests of statistical significance. Cohen’s d effect sizes were computed and interpreted based on the criterion of 0.2 (small effect), 0.5 (moderate effect), and 0.8 (large effect). For the outcome variables with non-normal distributions, we conducted the Wilcoxon signed-rank test, the non-parametric counterpart of a paired-sample t-test, using the ‘wilcox_test’ R package with alpha (α) at < .05 for two-sided tests. For non-parametric tests, effect sizes were calculated using the ‘wilcox_effsize’ R package, where r value is interpreted to be a small effect (0.10 – < 0.30), moderate effect (0.30 – < 0.50), and large effect (≥ 0.50).

## Results

### Feasibility and acceptability of the intervention

13 participants attended 4.6 days/week over 20 weeks in average (*min* = 3.6 days/week, *max* = 5.3 days/week; *SD* = 0.5 days/week). The CSQ-8 total score indicated that participants were highly satisfied with the intervention (*M* = 29.1, *SD* = 3.1). These results support the feasibility and acceptability of BMS-based multicomponent exercise training for 20 weeks in community-dwelling older adults with mild-to-moderate cognitive impairment.

### Cognitive functioning

Statistically significant changes were found in the MoCA total score (*t* = 4.71, *df* = 11, *p <* 0.001) with a large effect size (*d* = 1.36); the post-test score (*M* = 22.3, *SD* = 3.31) was higher than the pre-test score (*M* = 20.6, *SD* = 3.15). Ten out of 13 participants made improvements in the MoCA total score at post-test, whereas 2 participants scored the same with the pre-test, and 1 participant refused the test at post-test. However, no significant differences were found in the MoCA-MIS (*t* = -0.899, *df* = 11, *p =* 0.388). The results of the NIH Toolbox test indicated statistically significant changes in the Pattern Comparison Processing Speed test score (*t* = 2.83, *df* = 11, *p <* .05) with a large effect size (*d* = 0.816); the post-test score (*M* = 67.8, *SD* = 15.4) was higher than the pre-test score (*M* = 60.1, *SD* = 9.69). Ten out of 13 participants made improvements in the Pattern Comparison Processing Speed test score at post-test, whereas 2 participants scored lower than the pre-test, and 1 participant refused the test at post-test. Differences in the Flanker score did not reach significance (*t* = 1.81, *df* = 12, *p =* .098) although a moderate effect size (*d* = 0.522) was obtained; the post-test score (*M* = 77.5, *SD* = 12.7) was marginally higher than the pre-test score (*M* = 74.2, *SD* = 13). See Figure 1A – 1D for a summary of these results.

**Figure 1.**
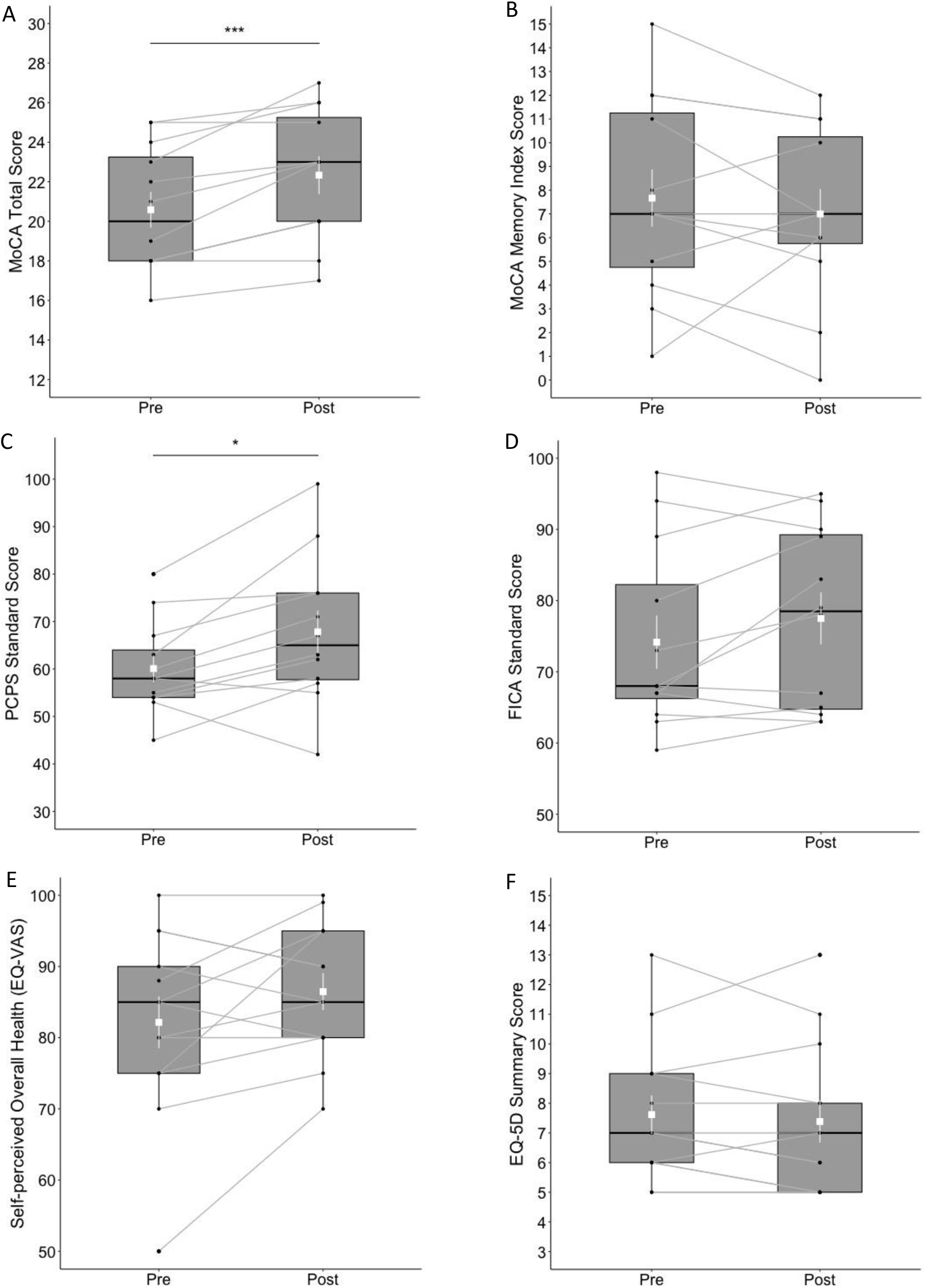
Box plots of cognitive and mental health outcomes at pre- and post-test; (A) MoCA total score, (B) MoCA Memory Index Score, (C) NIH Toolbox Pattern Comparison Processing Speed (PCPS) test, (D) NIH Toolbox Flanker Inhibitory Control and Attention (FICA) test, (E) self-perceived overall health reported on the EQ visual analogue scale (VAS), and (F) EQ-5D Summary Score. The shapes of the distribution are shown on the boxes and whiskers. The box bounds the IQR divided by the median (solid horizontal line) and whiskers extend to a maximum of 1.5 x IQR beyond the box. Mean and standard errors are indicated by small, white squares and appended lines. Significant differences between pre- and post-test are indicated by * p < .05, *** p < .001.

### Health-related QoL

No significant differences between pre- and post-test were found in the EQ-5D-5L summary score (*V* = 29, *p =* .273). The differences between pre- and post-test in EQ-VAS was not significant (*t* = 1.74, *df* = 12, *p =* .107); post-test score (*M* = 86.5, *SD* = 9.32) versus pre-test score (*M* = 82.2, *SD* = 13.1). See Figure 1E and 1F for a summary of these results.

### Physical functioning

The ranks of 6MWT total distance were significantly higher (*V* = 66, *p <* .05) at post-test (*Mdn* = 282 m) than at pre-test (*Mdn* = 249 m) with a large effect size (*r* = .601). Eleven out of 13 participants made an improvement in the 6MWT total distance. No significant differences between pre- and post-test were found in the ranks of TUG duration (*V* = 29, *p =* .273), 4SBT duration (*V* = 43, *p =* .398), and 30CST reps (*V* = 24.5, *p =* .798). See Figure 2 for a summary of these results.

**Figure 2.**
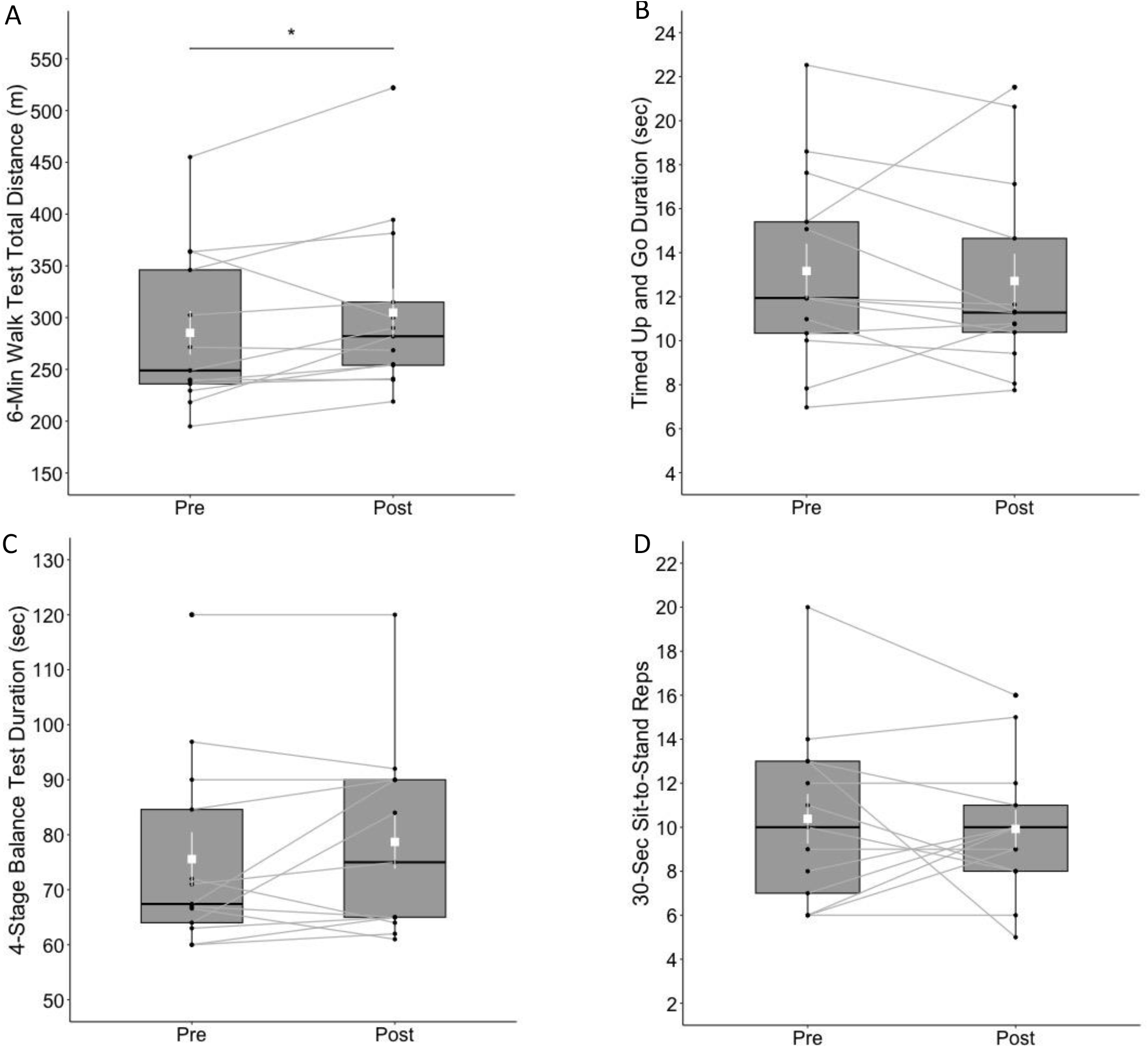
Box plots of physical functioning outcomes at pre- and post-test; (A) 6-Minute Walk Test total distance (m), (B) Timed Up and Go test, (C) 4-Stage Balance Test, and (D) 30-Second Chair Stand test. The shapes of distribution are shown on the boxes and whiskers. The box bounds the IQR divided by median (solid horizontal line) and whiskers extend to a maximum of 1.5 x IQR beyond the box. Mean and standard errors are indicated by small, white squares and appended lines. Significant differences between pre- and post-test are indicated by * p < .05.

## Discussion

Our participants demonstrated high adherence and satisfaction with the BMS-based multicomponent exercise intervention for 20 weeks. This finding supports the feasibility and acceptability of the intervention among older adults with mild-to-moderate cognitive impairment in an independent living facility. Our data also provide preliminary evidence in support of the efficacy of BMS-based multicomponent exercise training for cognitive and physical functioning in cognitively impaired older adults. Most participants who completed the intervention showed improvements in general cognitive functioning assessed by the MoCA, visual information processing assessed by the NIH Toolbox PCPS test, and walking endurance and aerobic fitness assessed by 6MWT. These findings are meaningful because we, for the first time, combined multicomponent exercise training with BMS and demonstrated its feasibility and preliminary efficacy for cognitive and physical health among cognitively impaired older adults.

It is possible that music stimulation played a positive role in participants’ high adherence and satisfaction with the exercise training. Scientists have demonstrated that listening to music prior to or during acute bouts of aerobic and resistance training have beneficial effects on affective valence, RPE, physical performance, and oxygen utilization, and thus becomes a motivational stimulant to PA bouts (for a review, see Terry et al., 2020). Despite this body of literature, there is an inadequate level of empirical evidence to substantiate the effects of music on long-term adherence to PA (Chair et al., 2021; Clark et al., 2022; Park et al., 2023). This gap in the literature was partly addressed by a theoretical model accounting for the putative mechanisms through which music acts to promote long-term adherence to PA (Park et al., 2023). From the view of the theory of hedonic motivation (Williams, 2018, 2019), music can help people like an exercise session more (or dislike it less) and thus increase wanting (or decrease dread) to exercise more or harder (Park et al., 2023). When people experience pleasure during an exercise session, this positive affective response is linked to enhanced motivation for another bout of exercise, which increases the chance of long-term adherence to PA (Park et al., 2023). Therefore, it is possible that participants in this study could benefit from positive affective responses to exercising with music. This is a tentative assertion at the moment because we did not measure affective response to PA nor did we include a control condition. Social interactions and the environmental factors in the independent living facility could have also made positive impacts on adherence. Future studies may investigate the unique effects of BMS on exercise adherence among older adults and psychological mechanisms underlying such effects.

Recent systematic reviews identified a few RCTs that demonstrated small but beneficial effects of music on long-term adherence to PA among older adults in a cardiac and pulmonary rehabilitation setting (Chair et al., 2021; Clark et al., 2022). The methodologies employed in this study, beat accentuation and tempo synchronization, may have played an important role in the observed high adherence rates given that music had little effect on long-term PA behaviors in RCTs without such methodologies. Specifically, older adults in a cardiac rehabilitation who were prescribed to walk-for-exercise with *beat-unaccented, tempo-asynchronous* music stimuli demonstrated trivial differences in the rate of meeting the PAG and accelerometer-measured PA over 26 weeks compared with controls who received the same exercise prescription without music (Clark et al., 2017). In another RCT, people with COPD who received an 8-week walking intervention with beat-unaccented, tempo-asynchronous music stimuli showed little difference in pedometer-measured and self-reported PA compared with controls in the same intervention without music (Bauldoff et al., 2002). However, in the RCT by Alter et al. (2015), walking-for-exercise with BMS led to nearly twofold increases in accelerometer-measured PA and in caloric expenditure over 3 months among midlife-to-older adults relative to the same exercise program with beat-unaccented, tempo-synchronous music and without music. This view would be supported by prior evidence that beat accentuation facilitates beat perception and auditory-motor synchronization when moving with music (Burger et al., 2013; Chen et al., 2006).

It is promising that BMS-based multicomponent exercise training for 20 weeks led to improvements in general cognition, cognitive processing speed, and walking endurance/aerobic fitness in cognitively impaired older adults. These findings are consistent with previous findings that exercise training has beneficial effects on cognitive functioning among older adults with normal cognition (Kelly et al., 2014; Northey et al., 2018), self-reported memory complaints (Barnes et al., 2013; Boa Sorte Silva et al., 2020; Lautenschlager et al., 2008), mild cognitive impairment (Baker et al., 2010; Wang et al., 2020; Zheng et al., 2016), and dementia (Groot et al., 2016; Wang et al., 2020). Moreover, the multicomponent nature of exercise training may have played a role for the positive outcomes in this study and thus has implications for future interventions to help older adults comply with the PAG. Our approach to prevent or delay the onset of AD or other dementias through multicomponent exercise training would be particularly urgent among older adults in early stages of cognitive decline.

Given the low rate of PA among older adults and the strong association between low PA and the prevalence of AD, developing and implementing a novel PA intervention for older adults will have implications for dementia prevention. This preliminary study will set the stage for an RCT to fully test the efficacy of BMS-based exercise training in the growing aging population at risk of dementia.

It should be noted that our intervention led to no changes in verbal memory (MoCA-MIS) and only nearly significant improvements in inhibitory control of attention (Flanker test). These findings are partly coherent with the meta-analytic findings that, in older adults with mild cognitive impairment, multicomponent exercise training has resulted in improvements in global cognition, attention, and executive function but not in memory (Wang et al., 2020) and aerobic training strongly improved global cognition but weakly improved memory (Zheng et al., 2016). We also attribute the lack of changes in the Flanker test to the lack of validity in some of our oldest-old participants at +90 years of age who had difficulties in understanding the test instructions and practice trials as well as the small sample size. This interpretation may be supported by the recent study which validated the NIH Toolbox Cognition Battery among healthy oldest older adults at 85–99 years of age who had MoCA total scores of 22–30 (Nolin et al., 2022), which is higher than the participants in this study. We also note that our intervention led to no changes in balance (TUG, 4SBT) and lower-body strength (30CST). We attribute these results to the limited capacity for balance training and lower body strength in the intervention. Due to the risks for falls and limited mobility, some oldest-older participants followed the entire exercises in a seated position, which minimized the training benefits for balance and lower body strength.

Limitations of this study are acknowledged. The small sample size and mild-to-moderate cognitive impairment of our participants and the convenient selection of an independent living facility might limit the ability to generalize these findings to the broader elderly population with and without cognitive decline. Furthermore, given the small sample size, we included all participants in the exercise intervention and were not able to include a no-exercise control group. Future studies may conduct an RCT to rigorously test the efficacy of BMS-based exercise training for physical, cognitive, and mental health among older adults with varying clinical conditions. Despite these limitations, the findings of this study are of value because they demonstrate the feasibility and acceptability of this intervention for cognitive impaired older adults and providing promising effect sizes that can be used to design future research.

### Clinical Implications

- Multicomponent exercise training can be beneficial for general cognition, cognitive processing speed, walking endurance, and aerobic fitness of older adults with mild-to-moderate cognitive decline in a community setting.
- Beat-accented music stimulation can be combined with exercise training to manipulate exercise tempo, which may be associated with good adherence to the training regimen by older adults.

**Table 1.** Multicomponent exercise intervention with beat-accentuated music stimulation

| Training goal | Dynamic warm-up | Aerobic step training | Muscle strengthening and balance training |  |  | Cool-down |
| --- | --- | --- | --- | --- | --- | --- |
|  |  |  | Upper body | Lower body | Core and balance |  |
| Frequency | 6 days/week | 6 days/week | 2 days/week (M, TH) | 2 days/week (T, F) | 2 days/week (W, S) | 6 days/week |
| Duration | 5 min | 15 min | 10 min | 10 min | 10 min | 5 min |
| Exercises | • Marching in place<br>• Elbow to knee taps<br>• Forward taps with punches<br>• Side taps with arm reaching | • Forward taps<br>• Backward taps<br>• Side taps<br>• Forward-backward steps<br>• Side steps<br>• Triangular steps<br>• Crossover steps | • Punches<br>• Bicep curls<br>• Bent over rows<br>• Shoulder presses<br>• Front arm raises<br>• Lateral arm raises | • Chair-assisted squats<br>• Lifting buttocks from the chair<br>• Straight leg raises - Front, Lateral, Rear<br>• Calf raises | • Chair-assisted deadlift<br>• Alternate knee raises<br>• Seated abdominal crunches<br>• Russian twists | • Trunk rotations<br>• Neck stretches<br>• Arm crosses<br>• Calf stretches<br>• Quadriceps stretches<br>• Hamstring stretches<br>• Gluteus maximus stretches |

*Note.* All exercises were performed in a chair-seated position or chair-assisted standing position. Participants were encouraged to exercise within light-to-moderate intensity by staying in 2-3 on the Ratings of Perceived Exertion (RPE) CR10 scale. M=Monday, T=Tuesday, W=Wednesday, TH=Thursday, F=Friday, S=Saturday

## Data Availability

All data produced in the present study are available upon reasonable request to the authors.

## Acknowledgement

This study was funded by the Undergraduate Research, Scholarship and Creativity Office (URSCO) and the Office of Leadership and Civic Engagement (OLCE) at the University of North Carolina at Greensboro. We sincerely thank to Gina Rice, Matthew Ward, and Csilla Roper, the staff members of the wellness team at the Heritage Greens Senior Living Community, for their support and cooperation for the implementation of intervention and testing in this study.

